# Determinants for detection of infection with SARS-CoV-2 Omicron variants of concern in health care workers by rapid antigen tests

**DOI:** 10.1101/2022.12.08.22283259

**Authors:** Jochen M. Wettengel, Katharina Strehle, Catharina von Lucke, Hedwig Roggendorf, Samuel D. Jeske, Catharina Christa, Otto Zelger, Bernhard Haller, Ulrike Protzer, Percy A. Knolle

## Abstract

**IMPORTANCE:** The rapid genetic evolution of SARS-CoV-2 and in particular the highly contagious Omicron variant of concern (VoC) may pose problems for rapid and accurate diagnosis of infection.

**OBJECTIVE:** Determine the diagnostic accuracy and robustness of a second generation rapid antigen tests compared to gold-standard, PCR-based diagnostics, for detection of infection with different SARS-CoV-2 Omicron VoC sub lineages in health care workers.

**DESIGN, SETTING, AND PARTICIPANTS:** The study included 428 health care workers from the University Hospital Munich Rechts der Isar of the Technical University of Munich who either reported recent onset of COVID-19 associated symptoms or completed routine diagnostic testing between 24^th^ of May and 22^nd^ of September 2022. All participants gave written informed consent to participate in this study and completed a questionnaire on infection-associated symptoms, prior SARS-CoV-2 infections and vaccination status.

**INTERVENTIONS:** During the first visit, two nasal swabs and one oropharyngeal swab were taken to perform two rapid antigen tests and a SARS-CoV-2 PCR-assay, respectively. A second set of nasal swabs was taken by the participants themselves two days later to repeat the two rapid antigen tests.

**MAIN OUTCOMES AND MEASURES:** The accuracy for detection of infection with different SARS-CoV-2 Omicron VoCs with two rapid antigen tests (*Test I* and *Test II*) was determined and compared to quantitative SARS-CoV-2 RNA levels detected by PCR.

**RESULTS:** In a side-by-side comparison, we found that *Test I* detected viral nucleocapsids from Omicron VoC (BA.5.2.3) at higher dilutions compared to *Test II*. In the 428 health care workers, *Test I* and Test II detected PCR-confirmed SARS-CoV-2 infection with different Omicron VoCs (BA.2, BA.4, BA.5) with a sensitivity of 89.4% (95% CI 81.9% - 94.6%) and 83.7% (95% CI 75.12% - 90.18%), respectively. Increased sensitivity of *Test I* was also reflected by earlier detection of SARS-CoV-2 infection. The lower test sensitivity of *Test II* could be compensated for by a repeated test performed two days later.

**CONCLUSIONS AND RELEVANCE:** Our study demonstrates that rapid antigen tests are suited to detect infection with the SARS-CoV-2 Omicron VoC and reveal an advantage of a lower detection limit for earlier detection of infection in health care workers.

## Introduction

The SARS-CoV-2 pandemic has led to more than 630 million infections and caused more than 6.6 million COVID-19 associated deaths. The rapid development of COVID-19 vaccines and their introduction into the clinic has helped to significantly reduce morbidity and mortality^1-5^. The evolution of SARS-CoV-2 and emergence of new variants of concern (VoC), however, poses a constant challenge^6,7^. Although being highly efficient in preventing severe COVID-19 disease courses, COVID-19 vaccination does not necessarily establish sterile anti-viral immunity ^8^. Emerging highly contagious variants, such as the Omicron VoC, carry mutations in their surface proteins enabling increased cellular receptor binding, attributing immune evasion properties or allowing for infection despite preexisting SARS-CoV-2 specific antibodies and T cells ^9-12^.

While COVID-19 vaccination provides protection from severe disease after SARS-CoV-2 infection in immune-competent vaccinated individuals, persons at risk because of pre-existing cardiovascular or lung disease, cancer patients or immune-suppressed patients can still develop sever disease^13^. Rapid detection of SARS-CoV-2 infection among health care workers is therefore important to limit infection transmission to patients at risk. While PCR-based diagnostics from oral swabs is the gold-standard for detection of SARS-CoV-2 infection, readily available and less costly rapid antigen tests allow self-testing and can reduce the time to diagnosis^14^. Rapid antigen tests have been extensively tested for their capacity to detect infection with SARS-CoV-2 VoCs^14-22^, which has shown a broad variability of the capacity to detect infections with SARS-CoV-2 VoCs.

We here conducted a diagnostic study to determine the value of a rapid antigen test with improved sensitivity for early detection of Omicron VoC SARS-CoV-2 infection in symptomatic health care workers in a university hospital setting.

## Methods

### Recruitment of participants

We aimed to compare the sensitivity of a first and a second generation rapid antigen tests for detection of infection with SARS-CoV-2 Omicron VoCs in health care workers. We therefore asked health care workers at the University Hospital München rechts der Isar who reported recent onset of COVID-19 associated symptoms or completed routine diagnostic testing between 24^th^ of May and 22^nd^ of September 2022 for their participation. All participants gave written informed consent to participate in this study. From 436 health care workers initially recruited, 428 participants completed all steps of the study (questionnaire, PRC testing, two rounds of rapid antigen tests) and were included into the study.

### Rapid antigen tests

Two rapid antigen tests (*Test I* – Roche SARS-CoV-2 Rapid Antigen Test 2.0 (9901-NCOV-09G) and *Test II* – SD Biosensor SARS-CoV-2 Rapid Antigen Test Nasal (9901-NCOV-03G)) were performed on the visit in the study center by the study team and two days later by the study participants themselves. For initial diagnosis, anterior nasal swabs were taken from all participants to perform the two rapid antigen tests and an oropharyngeal swab was taken for PCR-based diagnosis of SARS-CoV-2 infection. Two days later, participants repeated antigen testing by themselves. For these repeated tests, all participants received training for correct execution of anterior nasal swabs to assure comparability of results, and were randomized into two groups (group 1: *Test I* -> *Test II* or group 2: *Test II* -> *Test I*), which did not show any significant differences in test results. The results of this second set of tests were reported and documented via a photograph of the tests that was sent to the study team.

### PCR-based detection of SARS-CoV-2

For SARS-CoV-2 PCR analysis oropharyngeal swap samples were collected using Noble Bio REST CTM swabs. PCR-based detection of SARS-CoV-2 infection was performed in a routine diagnostics laboratory at the Institute of Virology on a Roche Cobas 6800 using the “Cobas SARS-CoV-2 test kit”, or on the Qiagen NeuMoDx using the “NEUMODX SARS-COV-2 ASSAY”. Quantification of the viral load was achieved by a normalized conversion equation using the ct value determined.

### SARS-CoV-2 genome sequencing

Genome sequencing of SARS-CoV-2 RNA was performed within the Bay-VOC network when viral loads determined by PCR were above 9.5×10^4^ GE/ml. RNA was extracted using a Seegene SeePrep32 device, and next generation sequencing was performed via Illumina next-generation sequencing to determine the SARS-CoV-2 pango lineage. SARS-CoV-2 genomes, which could not successfully be sequenced (18 out of 70 samples), are indicated as n/a.

### SARS-CoV-2 *in vitro* antigen assay

A clinical isolate SARS-CoV-2 Omicron swap sample (VoC BA.5.2.3, confirmed by viral genome sequencing) with a viral load of 1.9×10^8^ GE/ml was serially diluted in viral transport media. Subsequently, antigen test extraction buffer tubes were filled with 100 μl of the dilutions, vortexed and tests were performed according to the manufacturer‘s instructions.

### Ethics

The study was approved by the local ethics committee of the Technical University of Munich at the University Hospital München rechts der Isar (2022-265-S-Art.74-SR).

### Statistical analyses

Statistical analysis was performed using GraphPad Prism (GraphPad Software, San Diego, California USA) and R (R Foundation for Statistical Computing, Vienna, Austria). Means and standard deviations are presented for quantitative data, absolute and relative frequencies for categorical data. For calculation of sensitivity, specificity, positive predictive value and negative predictive value of the two rapid antigen tests, results of the PCR tests were considered as gold standard. Exact 95% confidence intervals (CIs) were estimated and are presented (Clopper-Pearson intervals). A logistic regression model was fit to the data to estimate the detection probability of both rapid antigen tests in dependence of the viral load using data from individuals with positive PCR tests. Results of the antigen tests were used as dependent variable (positive = 1, negative = 0) and the log-viral load as independent variable.

## Results

We compared a first and a second generation rapid antigen test for their capacity to detect infections with the SARS-CoV-2 Omicron VoC. Side-by-side comparison of the two rapid antigen tests using a serial dilution of a molecularly defined clinical isolate of the Omicron VoC BA.5.2.3 demonstrated that an 8-fold higher dilution of the clinical isolate were still detected by the second generation *Test I* compared to *Test II* (**Figure 1a,b**). We wondered whether the increased capacity to detect Omicron VoC nucleocapsids also translated into a more sensitive detection of infection with different SARS-CoV-2 Omicron VoCs in health care workers.

**Figure 1.**
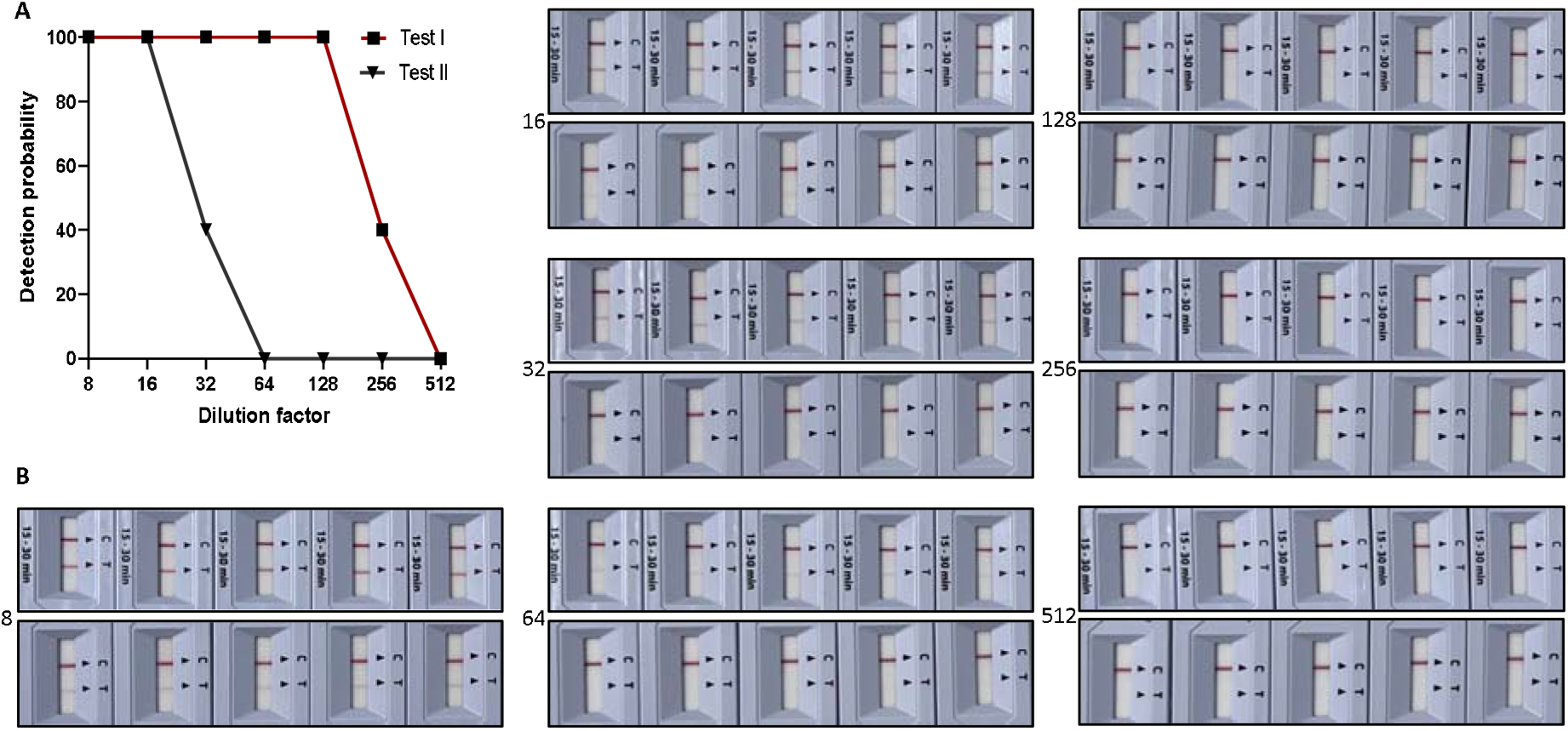
Sensitivity of two rapid antigen tests to detect nucleocapsids of a clinical isolate Omicron VoC (BA.5.2.3) **(A)** Serial dilution of a clinical isolate of the Omicron VoC of SARS-CoV-2 (BA.5.2.3) with 5 distinct measurements per dilution step, results were calculated for detection probability. **(B)** Images of the results of the rapid antigen *Test I* (upper row) and *Test II* (lower row), numbers on the left side denote the respective dilution factor.

We included 428 participants into the study who reported to the Coronavirus Diagnostic Center of the University Hospital München rechts der Isar because of COVID-19 associated symptoms or underwent routine testing in the absence of symptoms (**Table I**). Mean age of study participants was 36.3 years (range 18 – 67 years) with a similar age range in female (mean 34.4 years; range 18 – 67 years) and male (mean 37.0 years; range 19 – 64 years) participants (**Figure 2a** and **Table I**). All but 8 participants had received at least two COVID-19 mRNA vaccinations and 198 participants (46.3%) reported one or two prior SARS-CoV-2 infections (**Figure 2b, 2c, Table I**). 190 participants reported recent onset of COVID-19 associated symptoms, whereas 238 participants were asymptomatic (**Figure 2d, 2e** and **Table I**).

**Table I.** Characteristics of study participants.

| Characteristics | Participants (n = 428) |
| --- | --- |
| Sex |  |
| Female | 313 |
| Male | 115 |
| Age |  |
| All Mean (SD) | 36.33 (11.29) |
| Female Mean (SD) | 34.41 (9.33) |
| Male Mean (SD) | 37.04 (11.87) |
| SARS-CoV-2 vaccination, doses |  |
| #0 | 3 |
| #1 | 5 |
| #2 | 39 |
| #3 | 334 |
| #4 | 44 |
| #5 | 3 |
| Previous SARS-CoV-2 infections |  |
| #0 | 230 |
| #1 | 182 |
| #2 | 16 |
| Presenting with COVID-19 typical symptoms |  |
| No | 238 |
| Yes | 190 |
| Symptoms |  |
| Fever | 41 |
| Headache | 107 |
| Cough | 111 |
| Sore throat | 151 |
| Distress | 20 |
| Cold | 113 |
| Loss of taste/olfaction | 16 |
| Dorsalgia/joint pain | 68 |
| Exhaustion | 105 |
| Others | 18 |
| Contact with a SARS-CoV-2 infected person |  |
| private | 59 |
| work | 62 |
| both | 1 |
| n/a | 306 |

**Figure 2.**
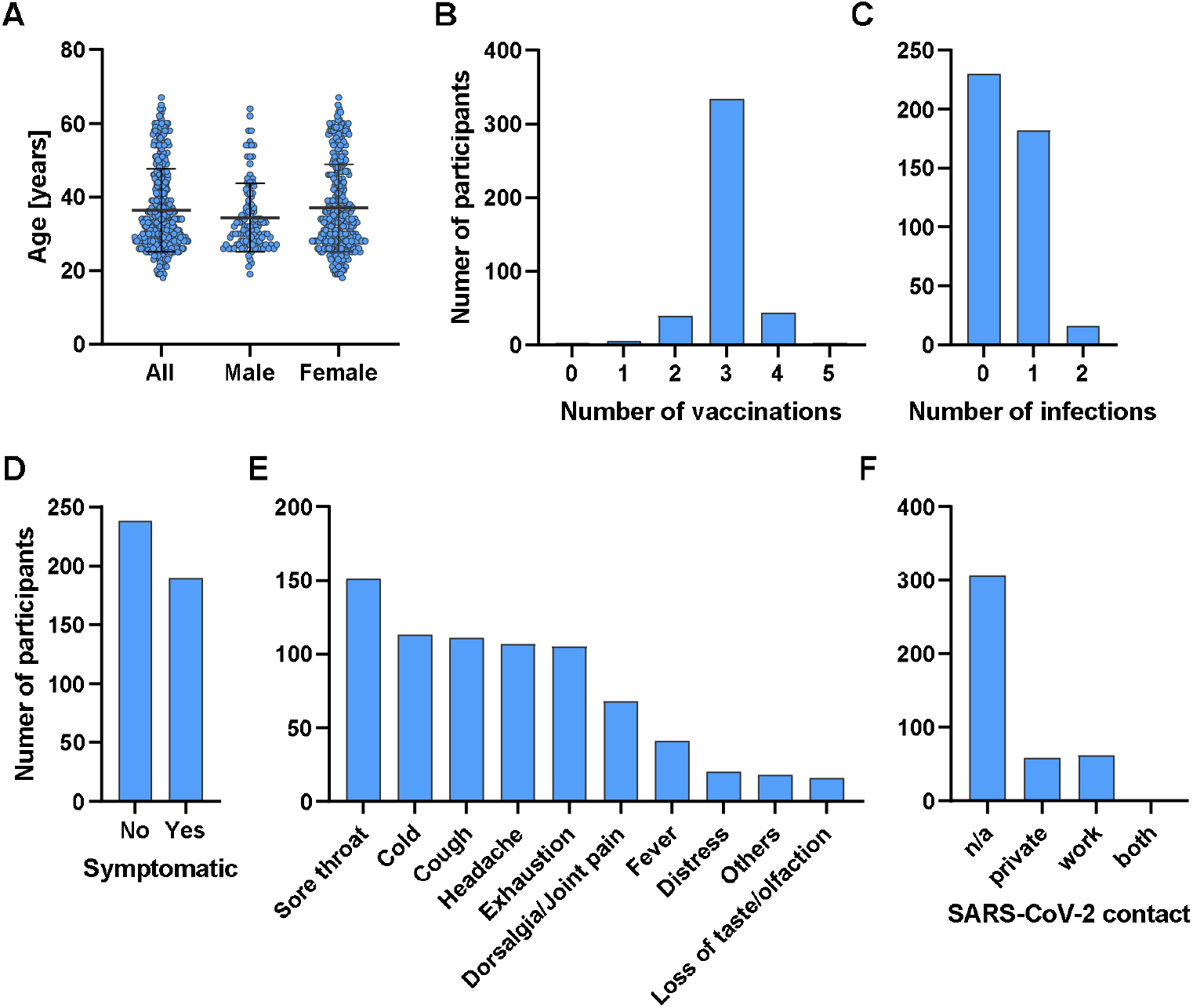
Characteristics of *RaCoMRI* study participants. **(A)** Age of participants enrolled in the *RaCoMRI*, bars indicate mean and standard deviations. **(B)** numbers of vaccinations received, or **(C)** numbers of prior SARS-CoV-2 infections. **(D)** Total numbers of participants reporting COVID-19 associated symptoms, **(E)** number of participants reporting specific symptoms, and (F) risk contacts with SARS-CoV-2 infected persons reported by study participants.

In 104 out of 428 participants included in this analysis we detected SARS-CoV-2 RNA by RT-qPCR (**Figure 3a**). CT values were used to determine virus load. Notably, seven patients were SARS-CoV-2 RNA PCR positive without reporting any symptoms, six of whom were also detected by both antigen tests. Results from quantitative SARS-CoV-2 PCR ranged from 4.4×10^2^ genome equivalents (GE)/ml to 1.4×10^9^ GE/ml with a mean of 2.4×10^7^ GE/ml. Omicron VoC sub lineage analysis of 52/70 samples with a viral load ≥ 9.5×10^4^ GE/ml by full viral genome sequencing revealed a majority of infections with BA.5 followed by BA.2 and BA.4 (**Figure 3b, 3c**,). No infections with other SARS-CoV-2 VoCs were determined (**Figure 3c**).

**Figure 3.**
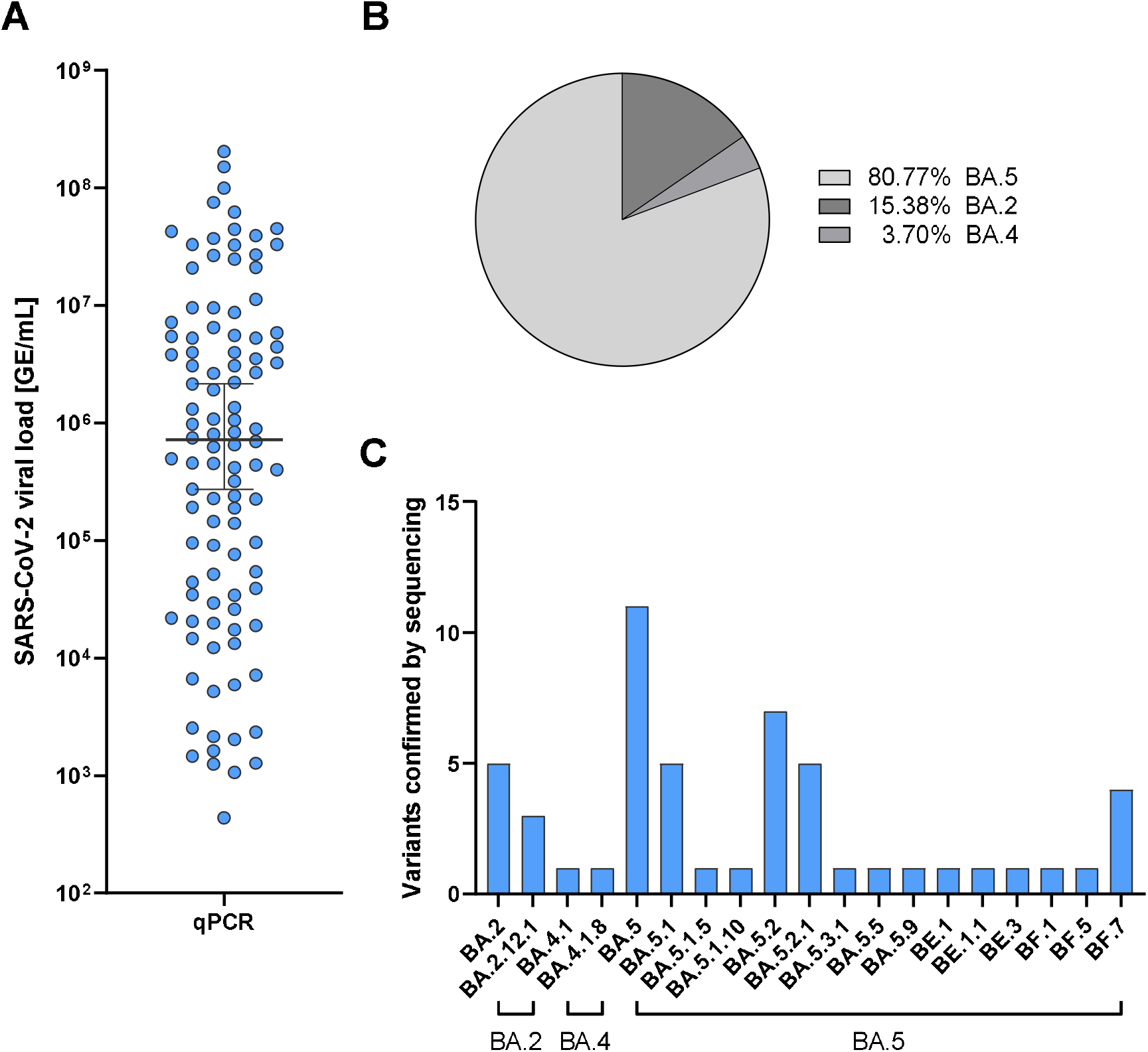
Quantitative results for SARS-CoV-2 RNA levels by PCR and results from viral genome sequencing. **(A)** Quantitative PCR results of SARS-CoV-2 RNA as genome equivalents (GE)/ml detected in oropharyngeal swabs from study participants. (B) Frequencies of Omicron VoC sublineages BA.5, BA.2 and BA.4 detected by full viral genome sequencing of samples with ≥ 9.5×10^4^ GE/ml. **(C)** Numbers of SARS-CoV-2 Omicron VoC pango lineages detected.

Using PCR-based results as reference, we detected 93 infections with Omicron VoCs with the second generation rapid antigen *Test I* (Table II). Eleven false negative results for *Test I* were noted, and two false positive results from 324 PCR-negative participants were obtained (**Figure 4a**). Overall, this resulted for *Test I* in a sensitivity of 89.4%, a specificity of 99.4%, a positive predictive value 97.9% and negative predictive value 96.7% (Table II). First generation *Test II* detected 87 infections among the 104 participants with PCR-confirmed SARS-CoV-2 infections, with 17 false negative results (**Table II**). For *Test II*, two false positive results from the 324 PCR-negative participants were obtained. This resulted for *Test II* in a sensitivity of 83.6%, a specificity of 99.4%, a positive predictive value of 97.8% and a negative predictive value of 95% (**Table II**). Side-by-side comparison of the false negative results with PCR results revealed that neither rapid antigen *Test I* nor *Test II* had an apparent cut-off level of viral RNA for detection of infection with both tests failing to detect some infections with low or high RNA levels of Omicron VoCs (**Figure 4**). Correlating the rapid antigen test results with the SARS-CoV-2 RNA levels that were detected by quantitative PCR revealed a higher detection probability for infection by *Test I* compared to *Test II* for infections with SARS-CoV-2 viral loads between 10^4^ to 10^6^ GE/ml (**Figure 5**).

**Table II.** Performance characteristics of the rapid antigen *Test I* and *Test II*.

|  |  | Test I |  |  | Test II |  |  |
| --- | --- | --- | --- | --- | --- | --- | --- |
|  |  | positive | negative | total | positive | negative | total |
| qPCR | positive | 93 | 11 | 104 | 87 | 17 | 104 |
|  | negative | 2 | 322 | 324 | 2 | 322 | 324 |
|  | total |  |  | 428 |  |  | 428 |
| Sensitivity (95% CI) |  | 89.42% (81.86% - 94.60%) |  |  | 83.65% (75.12% - 90.18%) |  |  |
| Specificity (95% CI) |  | 99.38% (97.79% - 99.93%) |  |  | 99.38% (97.79% - 99.93%) |  |  |
| Positive predictive value (95% CI) |  | 97.89% (92.60% - 99.74%) |  |  | 97.75% (92.12% - 99.73%) |  |  |
| Negative predictive value (95% CI) |  | 96.70% (94.17% - 98.34%) |  |  | 94.99% (92.09% - 97.05%) |  |  |

**Figure 4.**
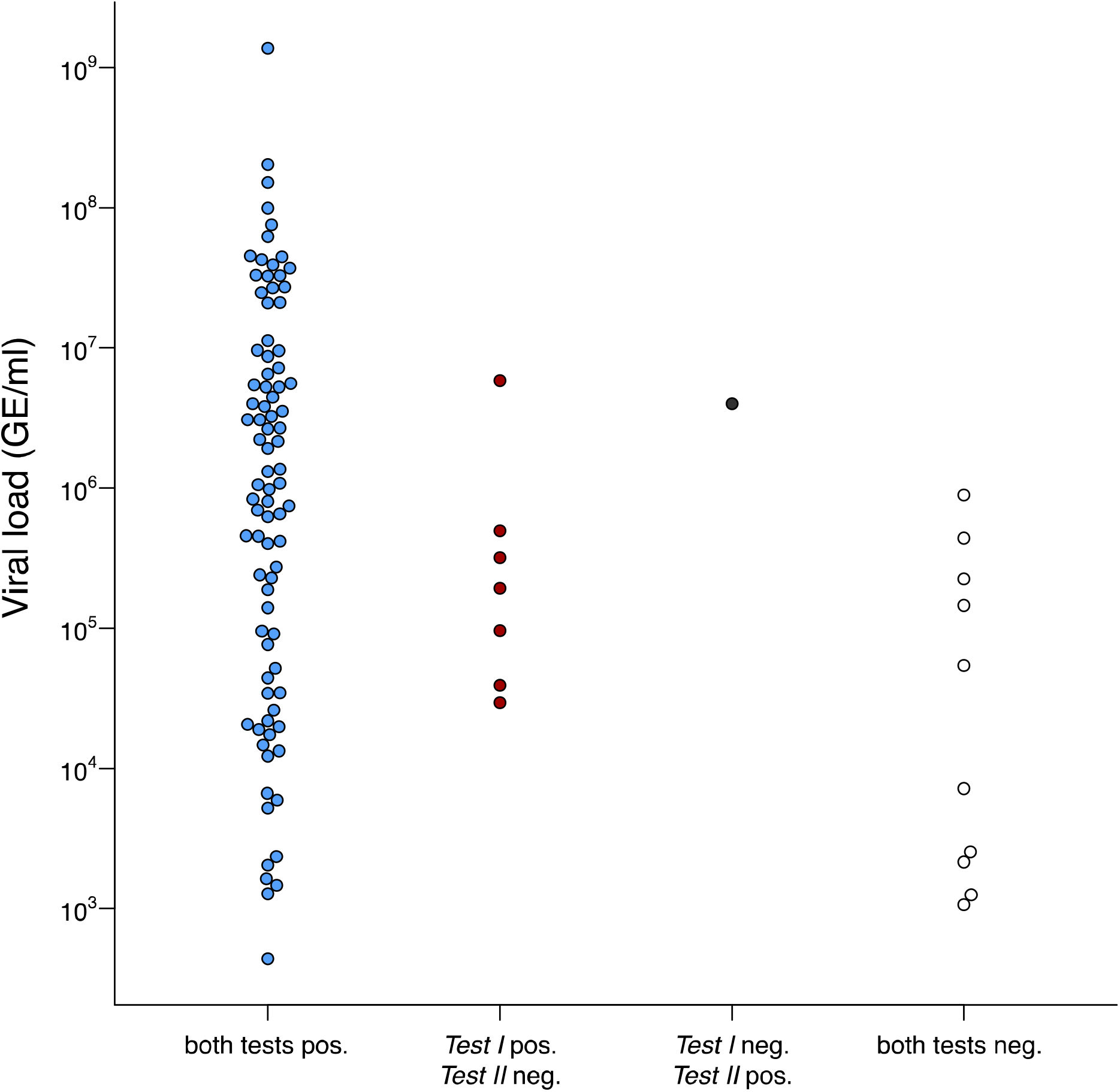
SARS-CoV-2 RNA levels related to test results obtained by *Test I* or *Test II*. Quantitative SARS-CoV-2 RNA levels from participants who had two positive rapid antigen tests (Test I and Test II positive, shown in blue), who were only positive by *Test I* (shown in red) or by *Test II* (shown in black), or who had to false negative tests (shown in open circles).

**Figure 5.**
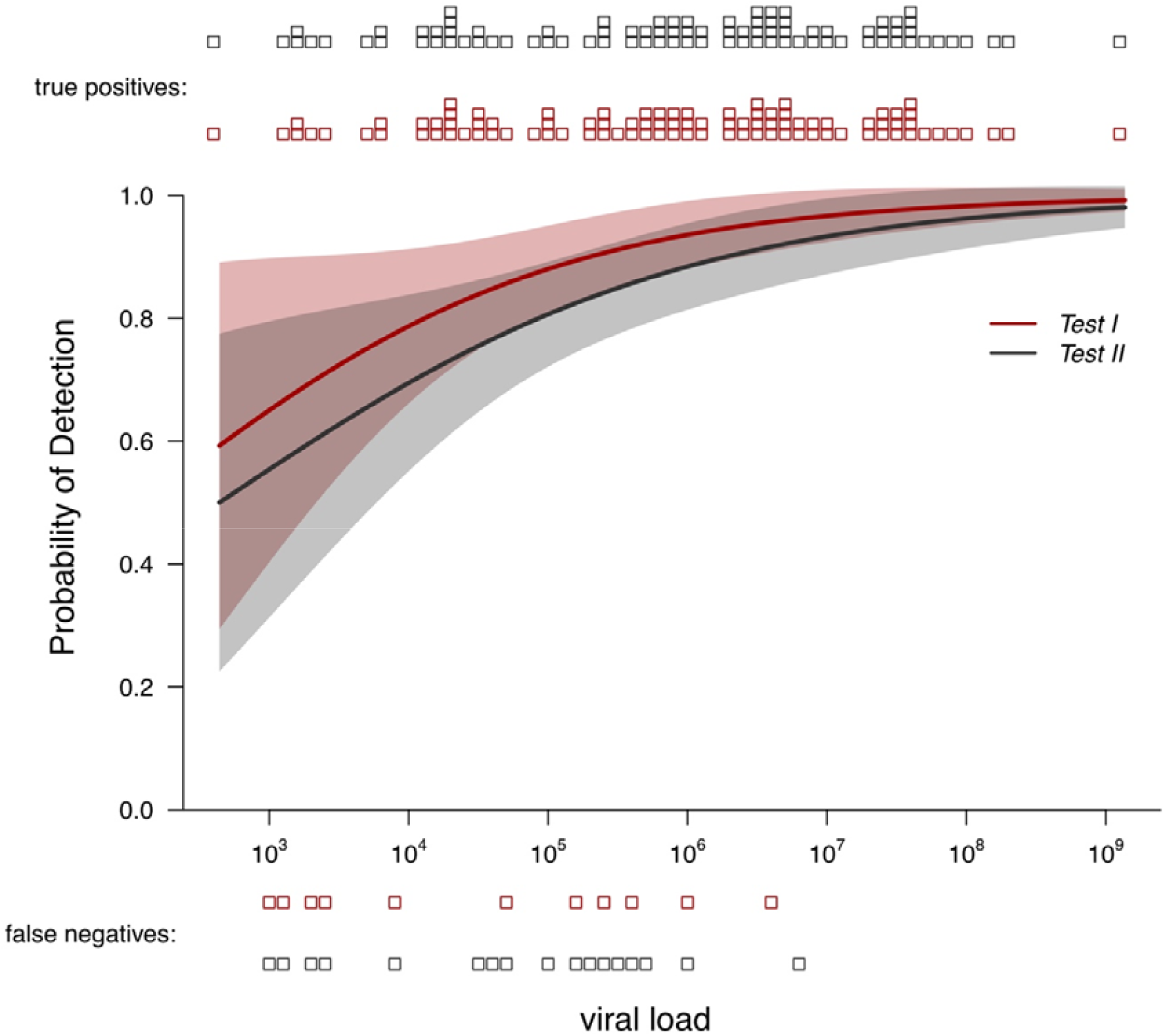
Detection probability for rapid antigen *Test I* and *Test II*. Detection probability by rapid antigen *Test I* and *Test II* stratified according to different SARS-CoV-2 RNA levels quantified by PCR.

Rapid antigen tests were repeated 48 hours later by the participants. When comparing the results of the rapid antigen *Test I* and *Test II* from the first with the second time point of analysis, we made several observations. First, in seven PCR-positive participants with a range of viral RNA levels from 2.9×10^3^ to 5.8×10^6^ GE/ml false negative results at the first time point were obtained by *Test II*, that were then correctly detected by *Test I*. At the second time point of analysis, rapid antigen *Test II* and *Test I* for these samples were both positive (**Table III**). Second, in five PCR-positive participants with viral RNA levels ranging from 7×10^3^ to 8.9×10^5^ GE/ml *Test I* and *Test II* both gave false negative results. At the second time point of analysis, both *Test I* and *Test II* gave positive results (**Table III**). Third, in four participants with very low viral RNA levels around 1-2×10^3^ GE/ml and one participant with 2.2×10^5^ GE/ml also repetition of the rapid antigen *Test I* and *Test II* two days later did not allow for correct detection of SARS-CoV-2 (**Table III**). Fourth, in two individuals with low viral RNA levels (4,3×10^2^ GE/ml and 5.9×10^3^ GE/ml) we detected positive results for rapid antigen *Test I* and *Test II* during the initial test but negative results for both tests two days later (**Table III**), most likely compatible with clearance of infection. Fifth, false positive results obtained from samples at the initial time point and two days later were consistent with *Test I* and *Test II* for samples from one participant giving consistently false positive results, for samples from one participant giving false positive results only at the initial time point and for samples from one participant giving false positive results only at the second time point (**Table III**). Combining the results from the two time points of analysis, *Test I* and *Test II* both detected 99 out of 104 SARS-CoV-2-infected PCR-positive participants with a sensitivity of 95.2% (**Table III**).

**Table III.**
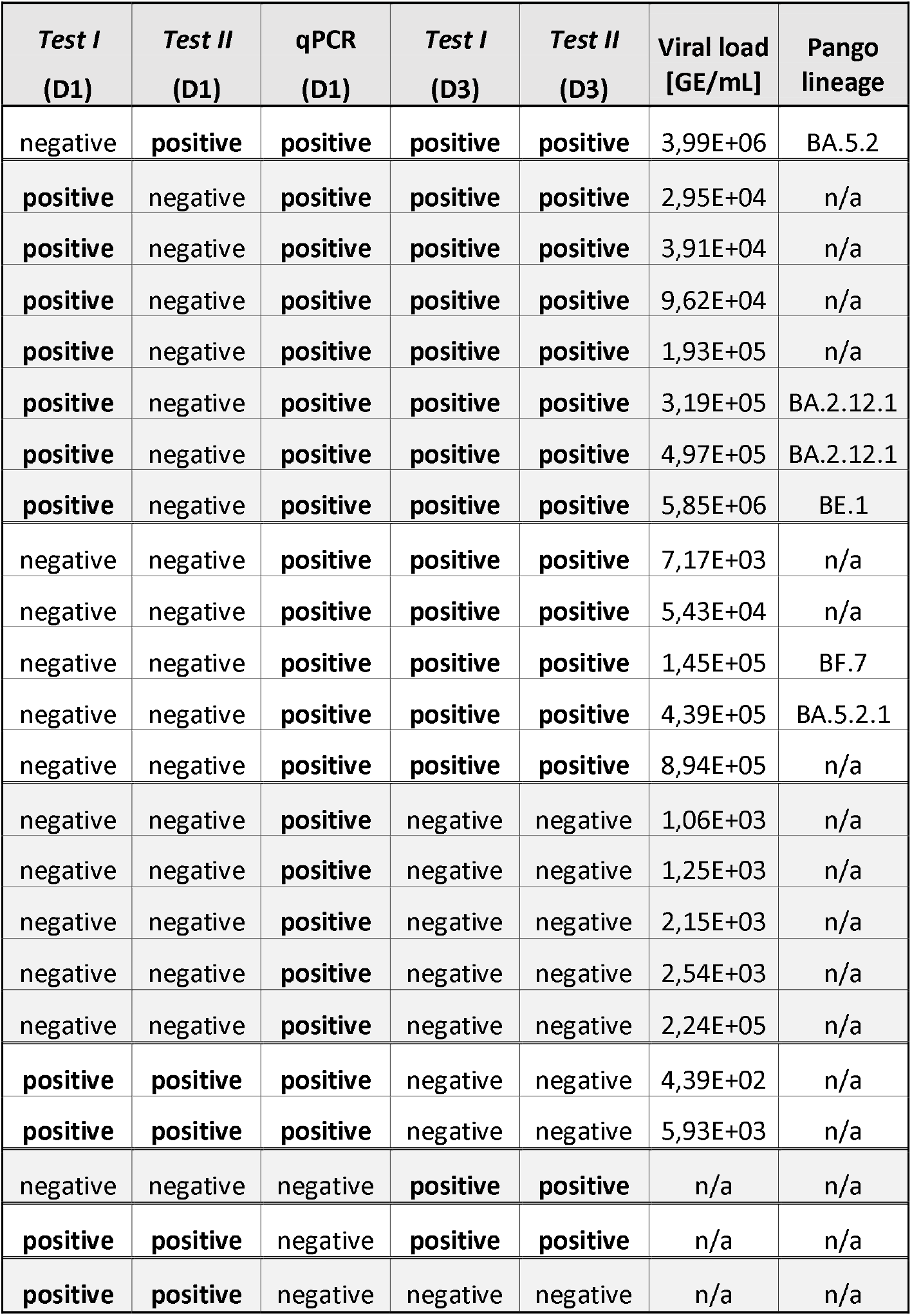
Results from the two time points of rapid antigen testing (Day1, Day3)

| *Test I*<br>(D1) | *Test II*<br>(D1) | qPCR<br>(D1) | *Test I*<br>(D3) | *Test II*<br>(D3) | Viral load<br>[GE/mL] | Pango<br>lineage |
| --- | --- | --- | --- | --- | --- | --- |
| negative | **positive** | **positive** | **positive** | **positive** | 3,99E+06 | BA.5.2 |
| **positive** | negative | **positive** | **positive** | **positive** | 2,95E+04 | n/a |
| **positive** | negative | **positive** | **positive** | **positive** | 3,91E+04 | n/a |
| **positive** | negative | **positive** | **positive** | **positive** | 9,62E+04 | n/a |
| **positive** | negative | **positive** | **positive** | **positive** | 1,93E+05 | n/a |
| **positive** | negative | **positive** | **positive** | **positive** | 3,19E+05 | BA.2.12.1 |
| **positive** | negative | **positive** | **positive** | **positive** | 4,97E+05 | BA.2.12.1 |
| **positive** | negative | **positive** | **positive** | **positive** | 5,85E+06 | BE.1 |
| negative | negative | **positive** | **positive** | **positive** | 7,17E+03 | n/a |
| negative | negative | **positive** | **positive** | **positive** | 5,43E+04 | n/a |
| negative | negative | **positive** | **positive** | **positive** | 1,45E+05 | BF.7 |
| negative | negative | **positive** | **positive** | **positive** | 4,39E+05 | BA.5.2.1 |
| negative | negative | **positive** | **positive** | **positive** | 8,94E+05 | n/a |
| negative | negative | **positive** | negative | negative | 1,06E+03 | n/a |
| negative | negative | **positive** | negative | negative | 1,25E+03 | n/a |
| negative | negative | **positive** | negative | negative | 2,15E+03 | n/a |
| negative | negative | **positive** | negative | negative | 2,54E+03 | n/a |
| negative | negative | **positive** | negative | negative | 2,24E+05 | n/a |
| **positive** | **positive** | **positive** | negative | negative | 4,39E+02 | n/a |
| **positive** | **positive** | **positive** | negative | negative | 5,93E+03 | n/a |
| negative | negative | negative | **positive** | **positive** | n/a | n/a |
| **positive** | **positive** | negative | **positive** | **positive** | n/a | n/a |
| **positive** | **positive** | negative | negative | negative | n/a | n/a |

## Discussion

The study results demonstrated that increased capacity for detection of SARS-CoV-2 Omicron nucleocapsids by a second generation rapid antigen test translates into an improved detection of infection with Omicron VoCs in health care workers. While other studies have demonstrated the ability of different rapid antigen tests to detect also infections with newly emerging Omicron sublineages^14^, this study for the first time reports on how a higher capacity for detection of viral nucleocapsids from emerging SARS-CoV-2 VoCs like Omicron translates into improved detection of infection with these VoCs in a cohort of health care workers independent of symptomatic infection.

Sensitivity of *Test II* under real-life conditions was improved compared to earlier results published ^23,24^ and met the 75% sensitivity required by regulatory agencies in Germany. No clear cut-off for SARS-CoV-2 RNA determined by quantitative PCR for detection of SARS-CoV-2 infection by both rapid antigen tests evaluated here could be defined. This points to other parameters relevant for accurate detection of infection such as extent of death of SARS-CoV-2 infected cells in the upper respiratory tract and associated release of viral nucleocapsids, that do not necessarily directly correlate with viral RNA levels. The results from this study will help to further improve strategies for early detection of SARS-CoV-2 infection with emerging VoCs like Omicron in symptomatic, but also in asymptomatic health care workers contributing to the prevention of spread of SARS-CoV-2 infection to vulnerable patient populations in hospitals.

### Strengths and Limitations

This study relies on the strength of combining a well-defined study cohort of health care workers infected with different SARS-CoV-2 Omicron sublineages and time-resolved analysis of infection by a first and a second generation rapid antigen tests. Taken together, the data support the notion that increased test sensitivity for detection of viral nucleocapsids translates into improved accuracy for detection of infection in particular at early time points. A further strength is the representation of different sublineages of SARS-CoV-2 Omicron detected in the infected study participants that reflect the broad spectrum of newly emerging variants. Limitations of the study are that the study period only spanned over 4 months and therefore did not include BA.1, BA.3, and newly emerging sublineages like BQ.1^25^.

## Conclusions

Our observation that the improved sensitivity for detection of viral nucleocapsids translates into increased detection accuracy for infection with the Omicron VoCs of SARS-CoV-2 lends support to the notion that a further increase of test sensitivity in the future is warranted to enhance the value of rapid antigen tests in accurate detection of SARS-CoV-2 infection. Our study confirms the limited sensitivity of rapid antigen tests and provides help in judging their role in test strategies at hospitals to detect infection in health care workers and prevent spread of SARS-CoV-2 infection to vulnerable patient populations.

## Data Availability

All data produced in the present work are contained in the manuscript

## Acknowledgments

We thank all the participants of this study, the study team and the diagnostic personal in the Institute of Virology at the TUM.

## Figures, legends and tables

**Supplementary Table I.**
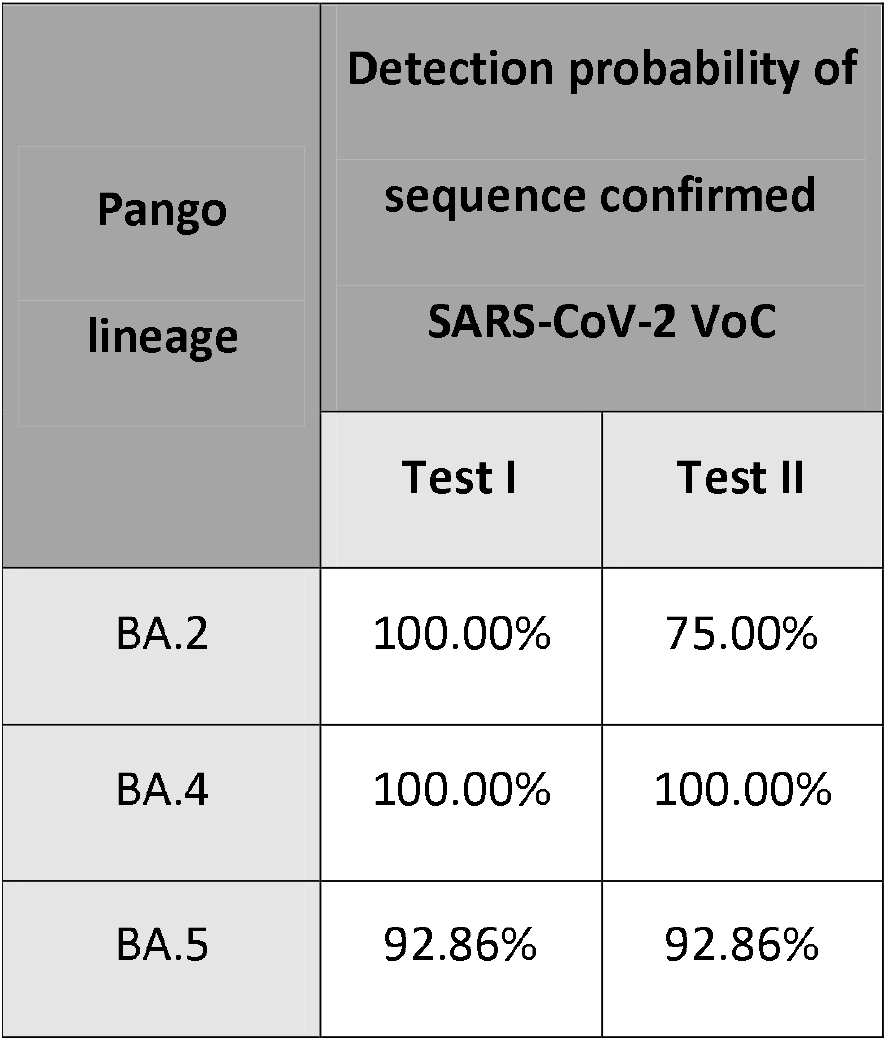
Detection probability of sequence confirmed SARS-CoV-2 VoC.

| Pango lineage | Detection probability of sequence confirmed SARS-CoV-2 VoC |  |
| --- | --- | --- |
|  | Test I | Test II |
| BA.2 | 100.00% | 75.00% |
| BA.4 | 100.00% | 100.00% |
| BA.5 | 92.86% | 92.86% |

**Supplementary Table II** | **All data collected in this study**

*See Excel Sheet*

## Notes

### Competing Interest Statement

The authors have declared no competing interest.

### Funding Statement

This study did not receive any funding. Reagents or direct assay costs were covered by Roche.

### Author Declarations

Ethics committee of the Technical University of Munich. Approval Number 2022-265-S-Art.74-SR

